# Autoimmune Encephalitis Related to Cancer Treatment with Immune Checkpoint Inhibitors: Systematic Review

**DOI:** 10.1101/2020.08.31.20185009

**Authors:** Vardan Nersesjan, Oskar McWilliam, Lars-Henrik Krarup, Daniel Kondziella

**Affiliations:** Department of Neurology, Rigshospitalet, Copenhagen University Hospital, Copenhagen, Denmark; Department of Clinical Medicine, Faculty of Health and Medical Sciences, University of Copenhagen, Copenhagen, Denmark

## Abstract

**BACKGROUND:** Immune checkpoint inhibitors (ICPI) are a game changer in the treatment of various metastasized cancers, but emerging reports of adverse events, including ICPI-associated autoimmune encephalitis (ICPI-AIE), are concerning. We aimed to collect all published cases of ICPI-AIE to identify the salient clinical and laboratory features of this disorder.

**METHODS:** We searched PubMed, The Cochrane Library and Embase for ICPI-AIE cases from the first description in 2015 until 01/2020 using standard bibliographic measures including PRISMA guidelines and pre-registration with PROSPERO (CRD42019139838).

**RESULTS:** Thirty-nine studies met inclusion criteria, resulting in 54 ICPI-AIE patients (mean age 58.6 years; 43% females). Common cancers included melanoma (30%) and non-small cell lung cancer (30%). Brain metastases were found in 16 patients (30%). The most frequent ICPI was nivolumab (61%). Onset of ICPI-AIE occurred on average after 3.5 treatment cycles, but very early and late presentations were common. Non-limbic AIE was roughly twice as frequent as limbic AIE (p<0.05). The most common laboratory abnormalities included bitemporal FLAIR lesions on MRI, continuous slow waves and diffuse slowing on EEG, and monocytic pleocytosis on cerebrospinal fluid analysis. Of note, intraneuronal antibodies were more frequent than neuronal surface antibodies, and logistic regression identified the presence of intracellular antibodies as a significant predictor for lack of improvement after 1^st^ line immunotherapy (p<0.05).

**CONCLUSIONS:** ICPI-AIE consists of a heterogenous group of conditions. Neurologists will likely encounter ICPI-AIE more often in the future, but important unresolved questions include the exact pathophysiological mechanisms, the epidemiology and the best treatment approaches associated with ICPI-AIE.

## INTRODUCTION

Immune checkpoint inhibitors (ICPI) have revolutionized the treatment of oncological patients. They are approved by the US Food and Drug Administration and the European Medicines Agency for the treatment of various advanced cancers like unresectable malignant melanoma, metastasized non-small cell lung cancer, and triple negative breast cancers.^1,2^ ICPI develop their anti-cancer properties by overriding a crucial mechanisms in the formation of metastatic cancer: Cancer cells usually evade immunosurveillance by activation of immune checkpoint pathways that lead to T lymphocytic Cell (TLC) apoptosis via several proteins, including cytotoxic T lymphocyte-associated Antigen 4 (CTLA-4), Programed Cell Death-1 protein (PD-1) and PD-1 Ligand (PD-L1). By inhibiting CTLA-4, PD-1 or PD-L1, ICPI reinstate the immune system’s antitumor response and promote immune-mediated tumor cell elimination.^1,3^

With the success of ICPI treatment, however, has followed evidence of immunological adverse events, notably autoimmune neurological adverse events (**Figure 1**).^4–6^ These events may affect both the central^4^ and the peripheral^6^ nervous systems. For instance, in 3,763 patients treated with ICPI, 1 % experienced neurological adverse events and 0.2 % developed autoimmune encephalitis.^4^ A pharmacological surveillance study of 48,653 treated with ICPI found that 0.51 % developed autoimmune encephalitis (AIE).^5^ Although a rare complication, ICPI-induced AIE is a potentially fatal condition and poses diagnostic challenges. The phenotype of ICPI-induced AIE may differ from classical limbic encephalitis or Anti-N-methyl D-aspartate (anti-NMDA) receptor encephalitis due to different disease-mechanisms and lack of detectable autoantibodies, and it may be challenging to distinguish between ICPI induced AIE and paraneoplastic induced AIE in cancer patients treated with ICPI.^3^

**Figure 1.**
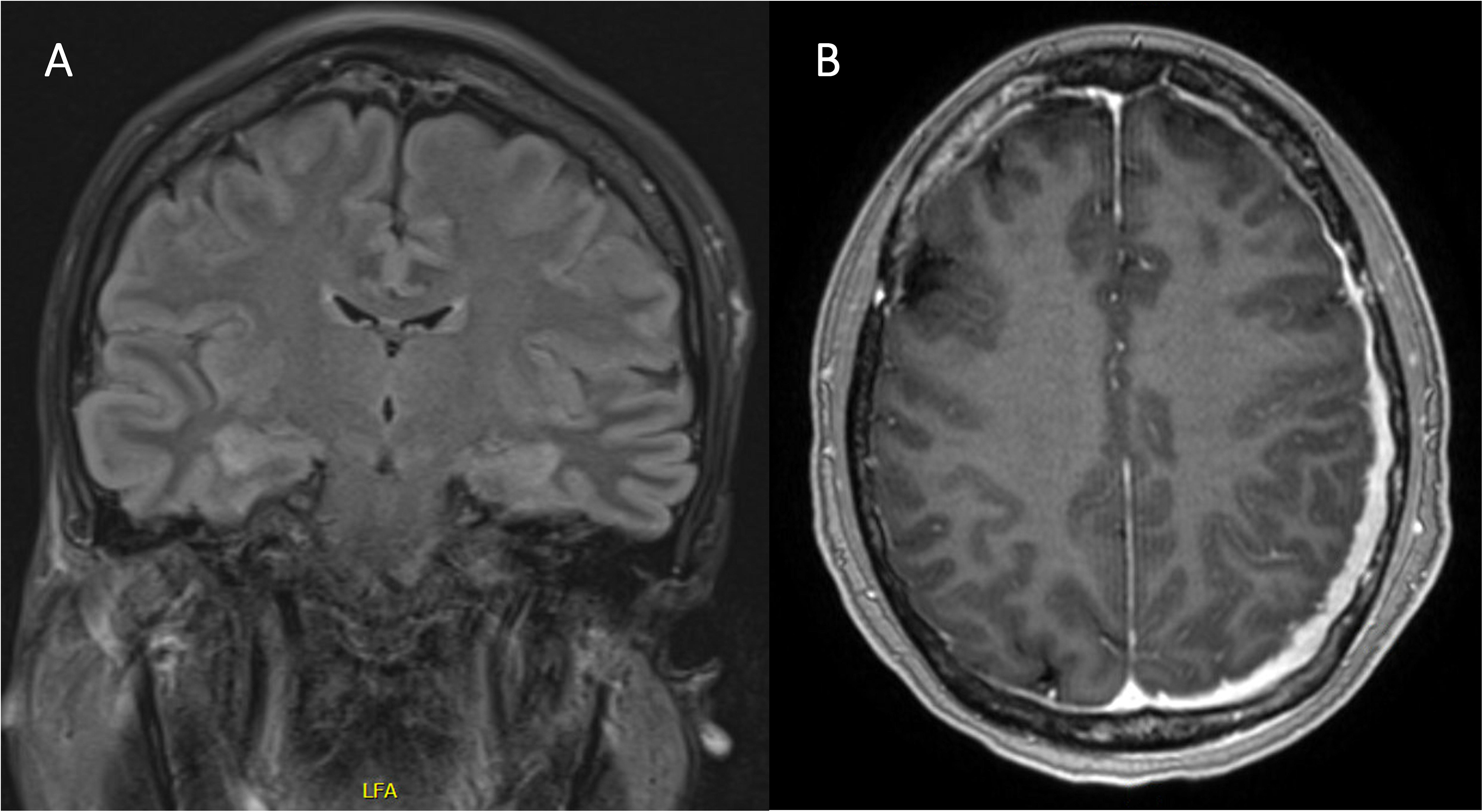
Written consent for publication of the case history was obtained from next of kin, but data have been removed on request from medRxiv for confidentiality reasons. However, the interested reader can contact the corresponding author for access to those data.

The rapidly increasing use of ICPI in antineoplastic treatment is destined to increase the frequency with which ICPI-associated AIE occurs. Several clinically important features remain poorly understood, however. It is unknown if ICPI-induced AIE is a clearly distinguishable entity in its own right or if it consists of a collection of heterogeneous conditions; if it has primarily a limbic presentation or if it involves extra-limbic areas as well; if neuronal surface or intracellular AIE antibodies are present or absent, and if the recently proposed AIE criteria^7^ are applicable to this disorder. We therefore performed a systematic review of all cases of ICPI-induced AIE published until January 2020 in order to characterize the salient symptoms, clinical findings, laboratory results and outcomes associated with the disease.

## METHODS

### Standard protocol approvals, registrations, and patient consent

We conducted a systematic review of the literature in accordance to the PRISMA guidelines.^8^ A PRISMA checklist was completed, and the review protocol was uploaded in PROSPERO (registration number CRD42019139838). Ethical approval was not required for this systematic review. The full protocol including search strings can be found at https://www.crd.york.ac.uk/prospero/display_record.php?RecordID=139838.

### Primary and secondary objectives

Using the patients, intervention, comparison, outcome (PICO^9^) approach, we phrased the following primary research question:

- In patients treated with ICPI (anti-PD-1, anti-PD-L1, and/or anti-CTLA-4) for disseminated malignancy, including systemic cancer, melanoma and hematologic malignancies, who develop rapidly progressive encephalopathy caused by brain inflammation in the absence of an infectious cause (P), are neurological examination and laboratory work-up including CSF analysis, EEG and brain MRI (I) compatible with a pure limbic encephalitis (C) or an encephalitis involving extra-limbic areas (O)?

We phrased two secondary research questions:

- In patients with ICPI-associated AIE, does plasma and cerebrospinal fluid (CSF) work-up for known autoimmune / paraneoplastic antibodies reveal the presence of those antibodies?
- Do patients diagnosed with ICPI-AIE during neurological examination and laboratory work-up, fulfill criteria for AIE as proposed by Graus et al.^7^?

### Search strategy

We evaluated all case reports, cross-sectional or longitudinal, retrospective or prospective observational studies as well as interventional trials reporting on patient history of autoimmune encephalitis following treatment with anti-PD-1, anti-PD-L1 and anti-CTLA-4 monoclonal antibodies.

We included only articles that allowed assessment of patient data at the single-subject level. We excluded articles that concerned patients already used in another article by the same authors (or the same institution). We included studies published in English and listed in Medline (PubMed), Cochrane Central Register of Controlled Trials (The Cochrane Library), and Embase since inception of these databases and up until January 2020.

We evaluated all abstracts and identified eligible studies based on full text review. We included only studies published in English, and reference list of each article was manually searched to identify additional articles. VN and OM performed initial selection and further review. After relevant studies were identified, VN and OM independently extracted relevant information. Disagreement was settled by LHK and DK. **Figure 2** visualizes data extraction points.

**Figure 2.**
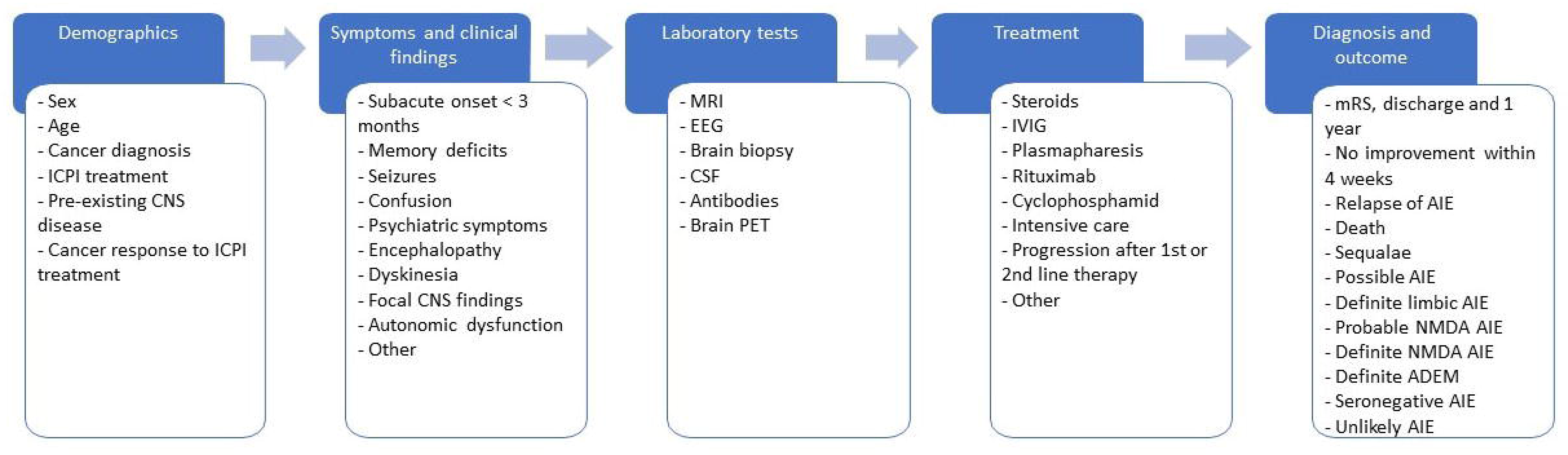
Overview of data points extracted from a systematic literature review

### Patients

We included adults (age ≥ 18 years) who were treated with anti-PD-1, anti-PDL-1 or anti-CTLA-4 for disseminated cancer and presented to a health care facility with symptoms suggestive of central nervous system (CNS) involvement and were diagnosed with encephalitis. Patients were included irrespective of co-morbidities, concomitant therapies and previous history of CNS disease.

### Target conditions

The target condition was AIE defined as a subacute onset of working memory deficits, altered mental status or psychiatric symptoms in combination with either seizure activity, CNS focal deficits, CSF pleocytosis or MRI features suggestive of encephalitis, in patients treated with nivolumab, pembrolizumab, atezolizumab, or ipilimumab as monotherapy or in combination. In line with previously proposed criteria for AIE by Graus et al.,^7^ we classified patients as having possible AIE, definite autoimmune limbic encephalitis, probable anti-NMDA receptor encephalitis, definite anti-NMDA receptor encephalitis, definite acute disseminated encephalomyelitis (ADEM) and probable autoantibody negative encephalitis.

### Statistical analysis

We used SPSS version 26 for statistical analysis. Fischer’s exact t-test was used to compare difference in dichotomized data, Mann-Whitney-U was used to analyze continuous data, and logistic regression was applied for outcome analysis of improvement after 1^st^ line therapy. p<0.05 was considered statistically significant.

## RESULTS

### A. Systematic literature search

The initial database searches retrieved a total of 603 studies, and 2 studies were manually added when checking reference lists (**Figure S1**, *supplemental files*). In total, 39 studies^10,11,20–29,12,30–39,13,40–48,14–19^ with 53 individual patients met inclusion criteria; the first study was published in 2015. We included data from the patient in **Figure 1**, seen at our institution. All studies were case reports or case series reporting on single subject data. No prospective data were available.

### B. Patient population

Of 54 patients included, 57 % were male and the mean age was 58.6 years. Eighty-nine % had metastasized cancer. The most common cancer diagnoses were melanoma (30 %), non-small cell lung cancer (NSCLC, 30 %) and renal cell carcinoma (7 %). Brain metastases were found in 16 patients (30%). The most common ICPI treatment was nivolumab (61 %), and dual ICPI therapy was initiated in 14 patients (26%). Complete remission of cancer was seen in 5 patients (9%) after ICPI treatment, positive responses despite incomplete remission in 16 patients (30%), and in 12 patients (22%) there was progression of cancer despite ICPI treatment. The mean cycle of ICPI treatments before occurrence of symptoms of presumed AIE was 3.5 (median 3.0). Ten patients (19%) experienced other autoimmune adverse events such as hypophysitis, dermatological manifestations and neuromuscular affections.

We divided patients in the following (partly overlapping) subgroups: With brain metastases (BM+) (N=16)^11,13,17,23,28,31,33,35,45,47,38,41,42,44,46^, without brain metastases (BM-) (N=38)^10,12,24–27,29–32,34,36,14,37,39,40,42,43,45,46,48,15,16,18–22^, with limbic AIE (N=16)^14,18,46,47,19,22,26,27,34,36,39,42^ and extra-limbic AIE (N=38)^10,11,23–26,28–33,12,35,37,38,40–42,44–46,48,13,15–17,20–22^. Limbic AIE were determined according to criteria for AIE by Graus et al.^7^ One patient had bitemporal FLAIR lesions and anti-Ma2 antibodies present in CSF, but the clinical presentation was not consistent with involvement of the limbic system, there were no CSF pleocytosis or EEG activity suggestive of temporal involvement, and this patient was therefore labelled as having extra-limbic AIE.^46^

Theses subgroups (i.e. BM+ vs. BM-and limbic AIE vs. extra-limbic AIE) were compared for differences in demographics, but there were no statistically significant differences. See **Table 1** for demographics.

**Table 1.**
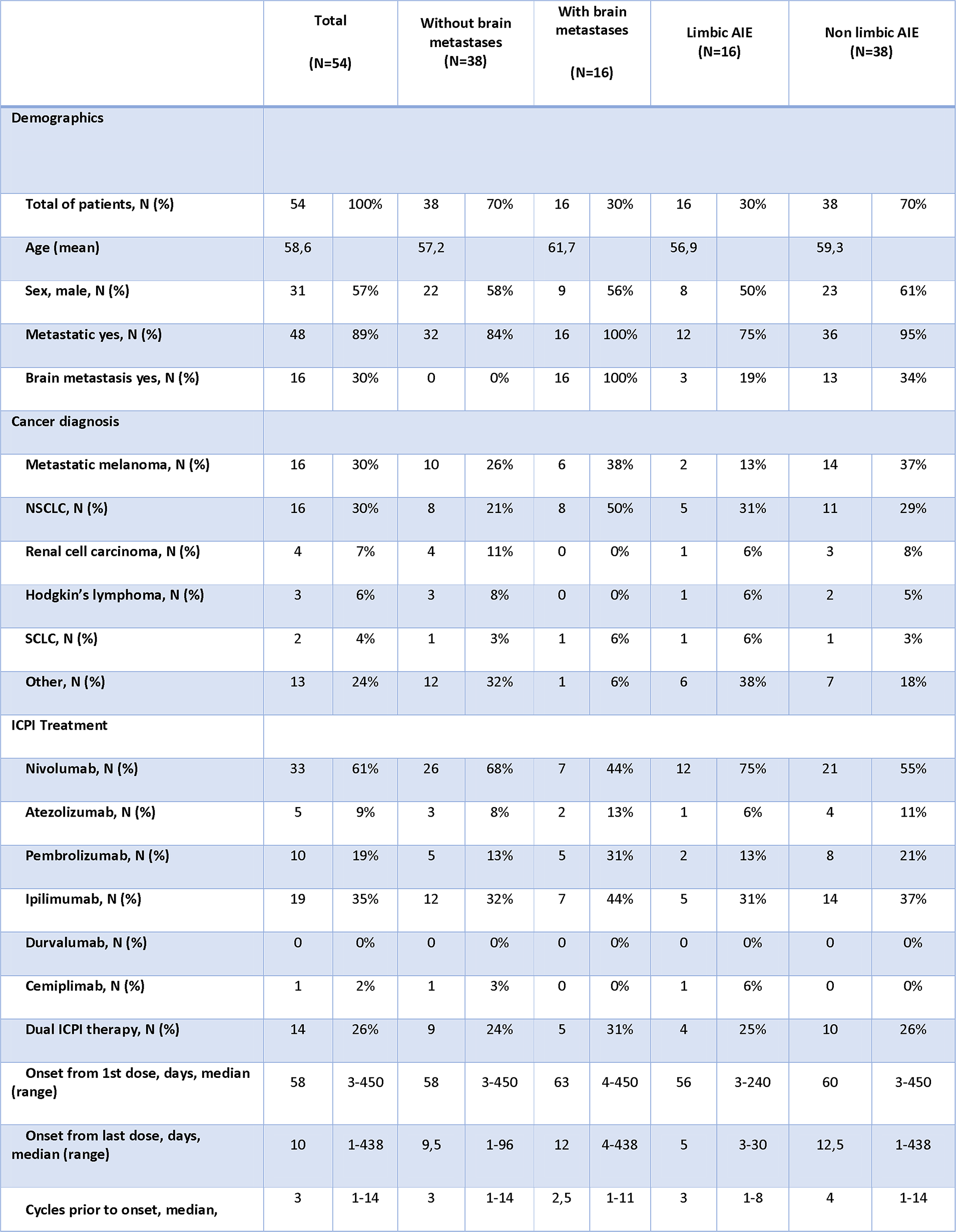
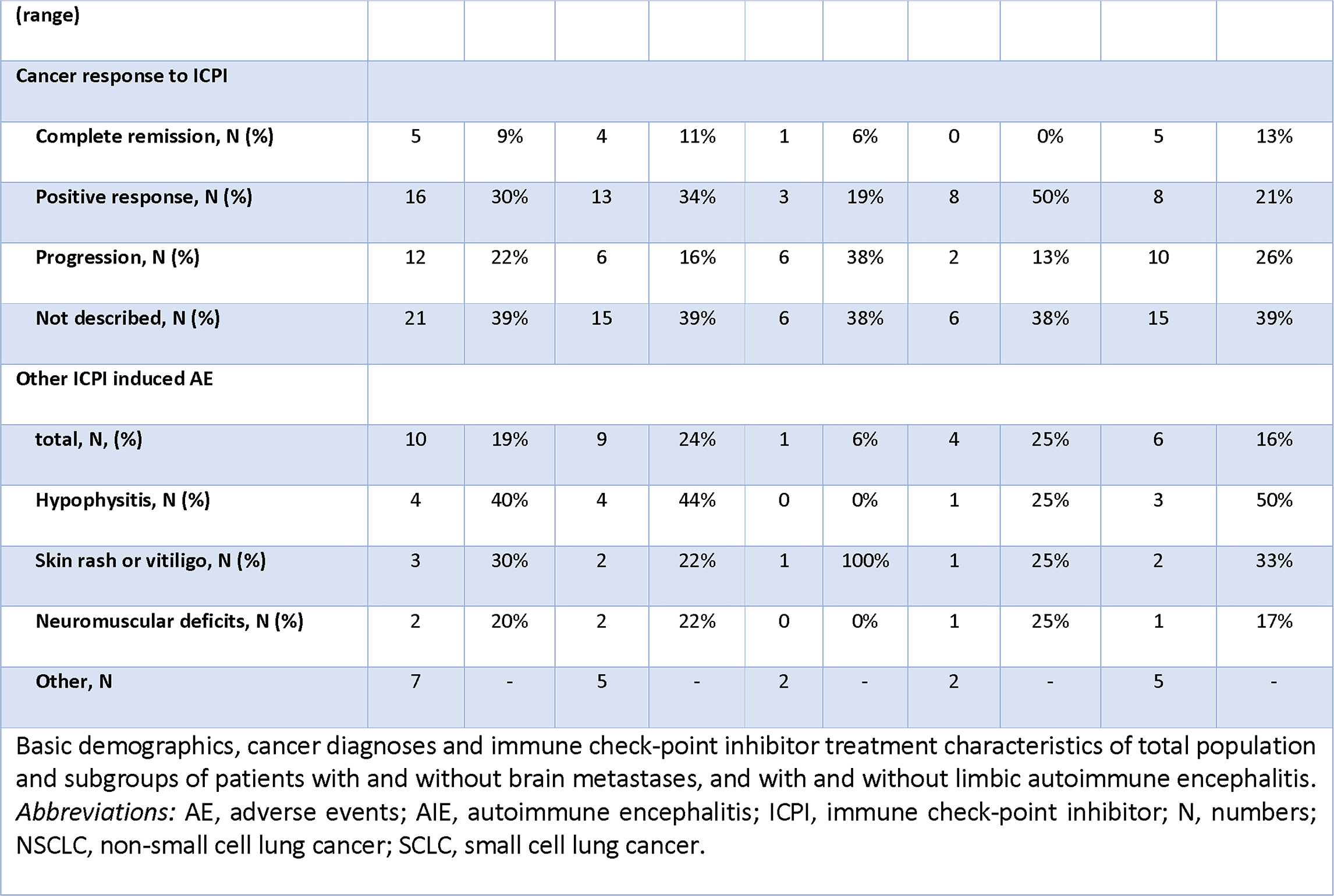

### C. Clinical symptomatology

**Table 2** shows an overview of symptoms from the total population and the subgroups. Symptom onset was subacute (< 3 months) in all patients^23^ except in one patient who had a sudden onset of symptoms, two patients^39,46^ with slow and progressive course, and one patient^13^ in whom symptom onset was not disclosed. Symptom and signs included, in decreasing frequency, altered mental status (85%), focal CNS deficits (63%), psychiatric symptoms (37%), seizures (33%), autonomic dysfunction (33%), working memory deficits (28 %), ataxia (19 %) and dyskinesia (11 %). Of the psychiatric symptoms, behavioral disorders were most frequent (55%), followed by affective symptoms (35%), hallucinations (20%) and paranoia (20 %). Concerning epileptic seizures, generalized seizures were most common (39%), followed by focal seizures (28 %) and non-convulsive status epilepticus (22 %). Eleven % had both generalized and focal seizures. Autonomic dysfunction was described in 13 patients, 2 (11%) of which had central hypoventilation and 12 (67%) of which had fever.

**Table 2.**
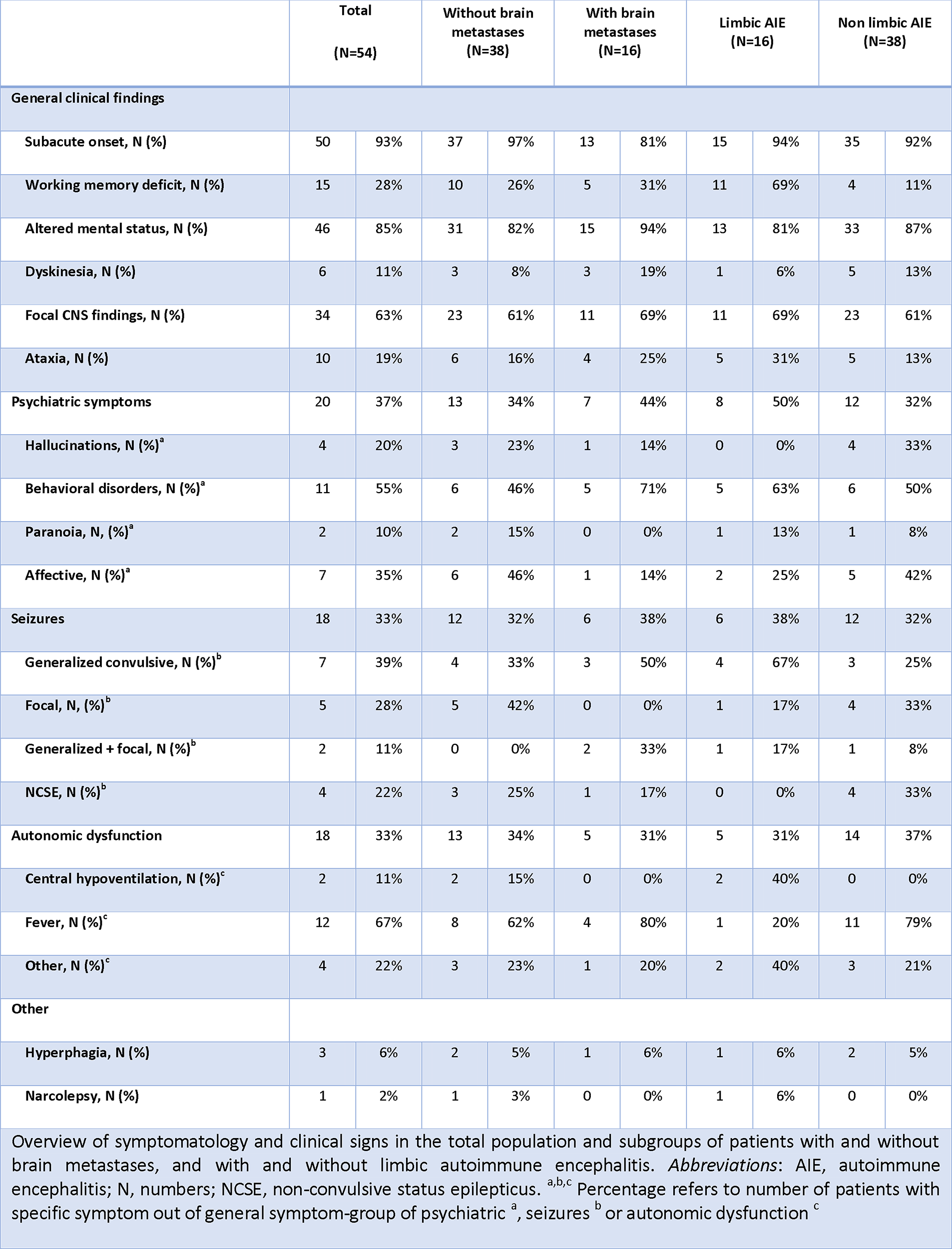

Working memory deficits were more frequent in patients with limbic AIE (N = 11, 69 %) than in patients with the extra-limbic AIE (N = 4, 11 %) p<0.001. No other symptoms occurred with statistically different frequency in the tested subgroups.

### D. Laboratory findings

#### Magnetic resonance imaging

Fifty-two patients (96 %) were investigated with MRI. 29 patients (56 %) had positive findings, with bitemporal FLAIR lesions (52%) being the most common followed by multifocal inflammatory lesions of white and/or grey matter (17%), leptomeningeal enhancement (14%), lesions of basal ganglia (7%) and pachy-meningeal enhancement (7%). One patient had a unilateral temporal FLAIR lesion^47^ and one patient had a not further specified lesion.^31^ MRI lesions were found in all patients with limbic-AIE, but only in 13 patients (36%) of the extra-limbic group (p<0.001). See **Table 3** for laboratory findings.

**Table 3.**
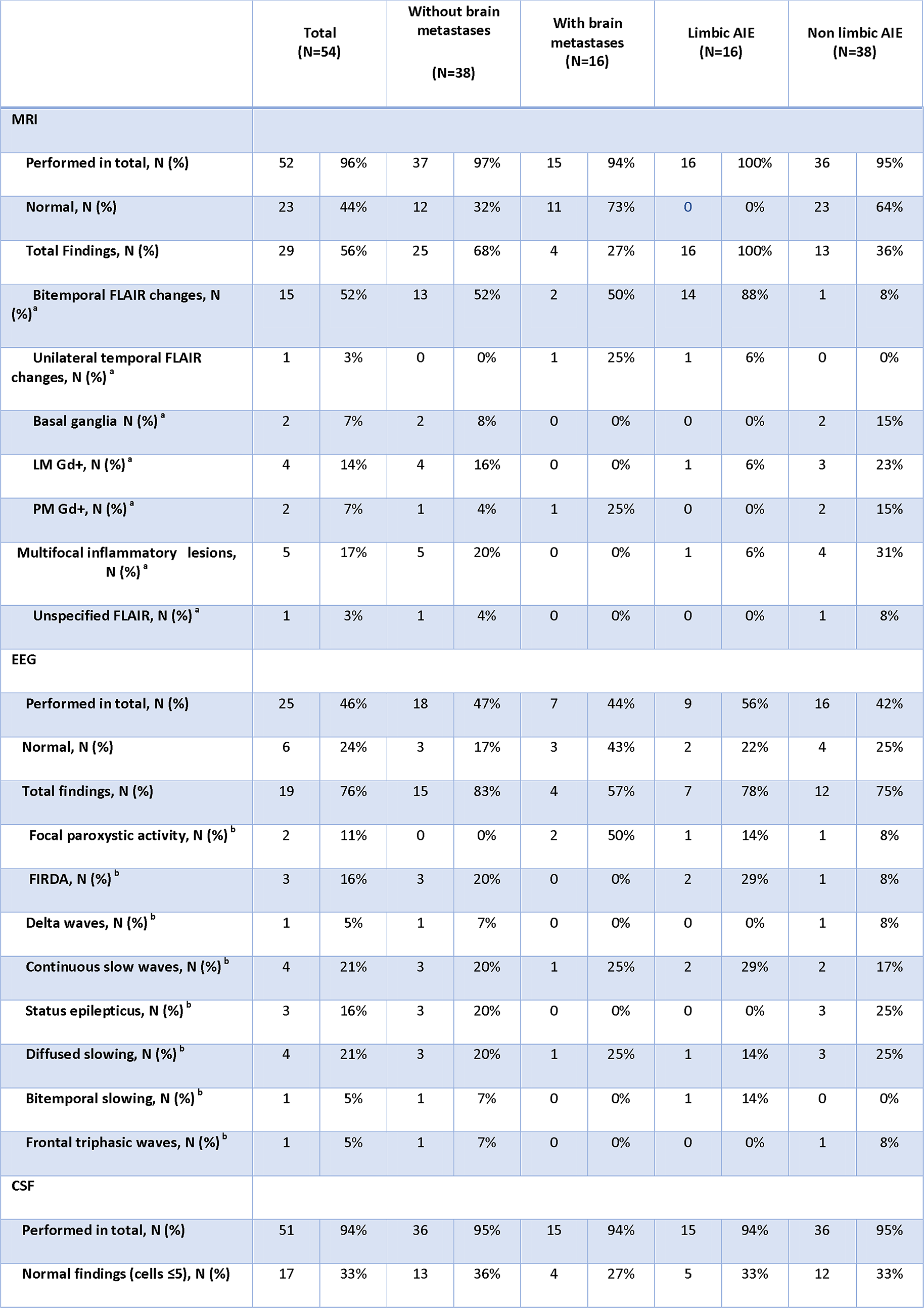
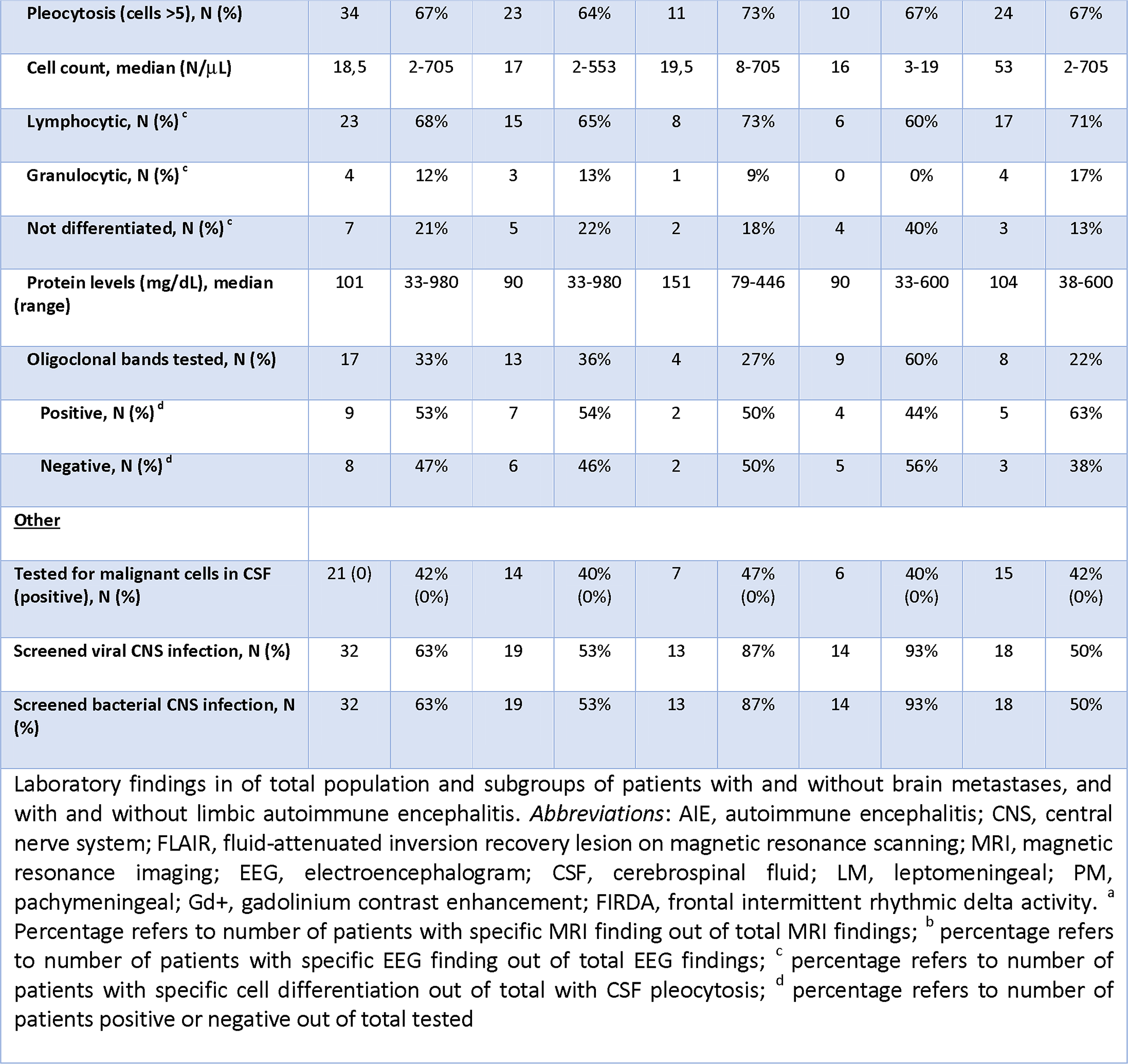

#### Electroencephalogram

EEG was performed in 25 (46 %) patients, of which 19 (76 %) had abnormal findings consisting of continuous slow waves (21 %), diffuse slowing (21 %), frontal intermittent delta activity (FIRDA) (16 %) and (non-convulsive) status epilepticus (16 %).

#### Lumbar puncture

CSF was analyzed in 51 patients (94 %) of which 34 patients (67 %) had pleocytosis (defined as >5 white blood cells/microL) with a median of 18 white blood cells/microL. Patients with limbic AIE had the same frequency of CSF pleocytosis but significantly fewer cells in the CSF compared to patients with extra-limbic AIE (median 16 vs. 53 white blood cells/microL, p<0.05). Cell differentiation in CSF was mostly lymphocytic (68 %). Four patients (12%) had predominantly granulocytes and all of these were in the extra-limbic group. Oligoclonal bands were searched for in 17 patients (33 %), and 53% of those were positive. Patients in the limbic AIE group were more frequently tested for oligoclonal bands compared to those from the extra-limbic AIE group (60% vs. 22 %, p<0.05), but there was no difference in frequency of positive oligoclonal bands between the groups.

**Table 4** shows an overview of antibody testing. Antibody testing for antibodies against cell-surface or intracellular antigens was performed in 57% (CSF) and 46% (blood) of patients, respectively. Of those tested, 55% and 48% had positive findings in CSF and blood, respectively. Antibodies against intracellular antigens in CSF were most frequent: anti-Ma2 antibodies occurred in eight patients^30,43,46^ (47%), anti-Hu in two patients^26,27^ (12%), anti-GAD in two patients^18,42^ (12%), an unspecified Purkinje cell antibody in one patient^27^ (6%) and anti-Ri in another one^26^ (6%). One patient^27^ had both anti-HU and an unspecified Purkinje cell antibody. Antibodies against cell-surface antigens were less frequent and only seen in three patients; anti-NMDA receptor^31,41^ and contactin-associated protein–like 2 (CASPR2) antibodies.^14^ One patient was described with a novel and unclassified antibody.^42^ Antibodies present in blood followed almost the same pattern of distribution in the population as CSF antibodies, although one patient had anti-glial nuclear antibodies in blood which was not present in CSF,^47^ one patient had positive anti-Hu present in blood before initiation of ICPI treatment,^34^ and one patient had elevated anti thyroid peroxidase antibodies (anti-TPO).^16^

**Table 4.**
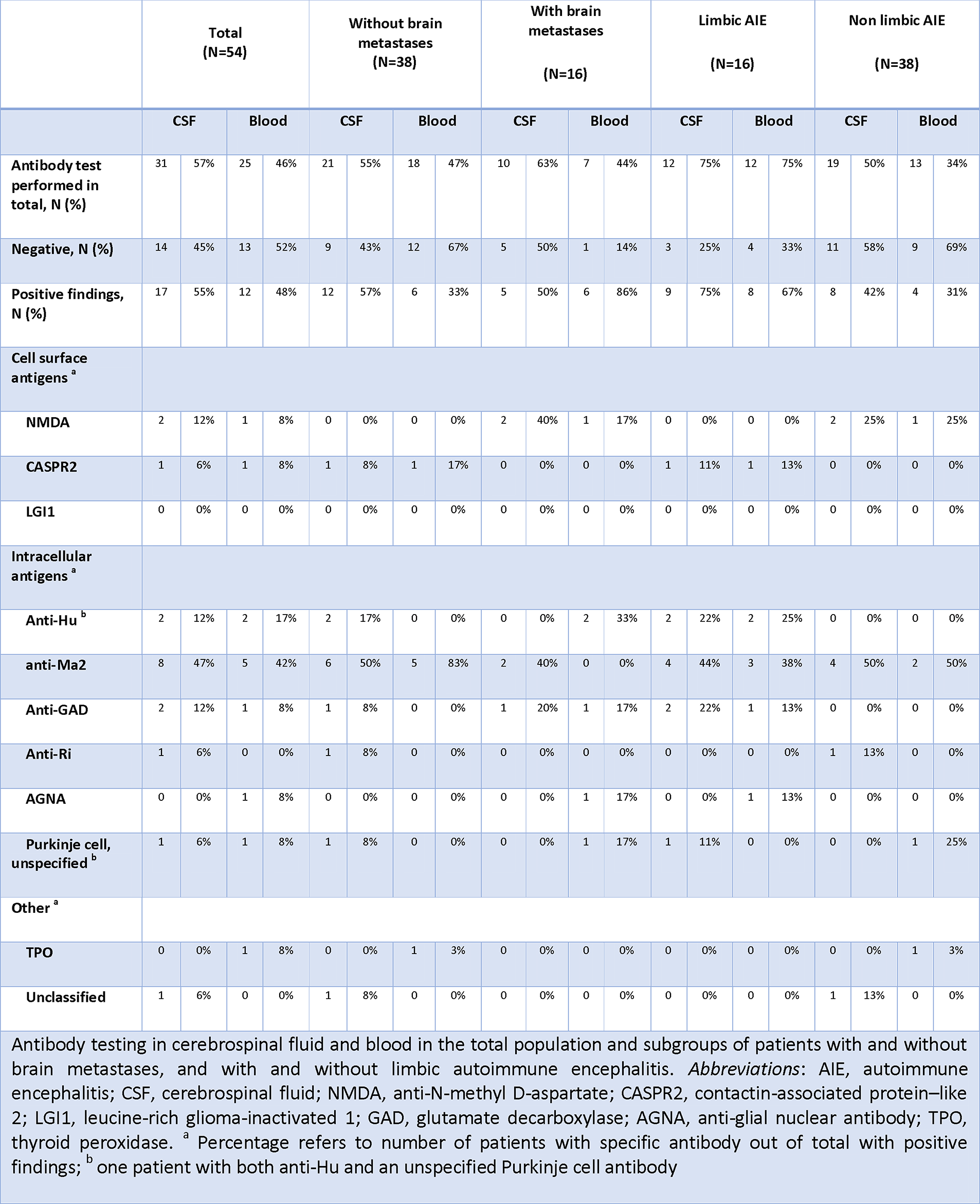

### E. Treatment and outcome

See **Table 5** for full outline of treatment and outcome.

**Table 5.**
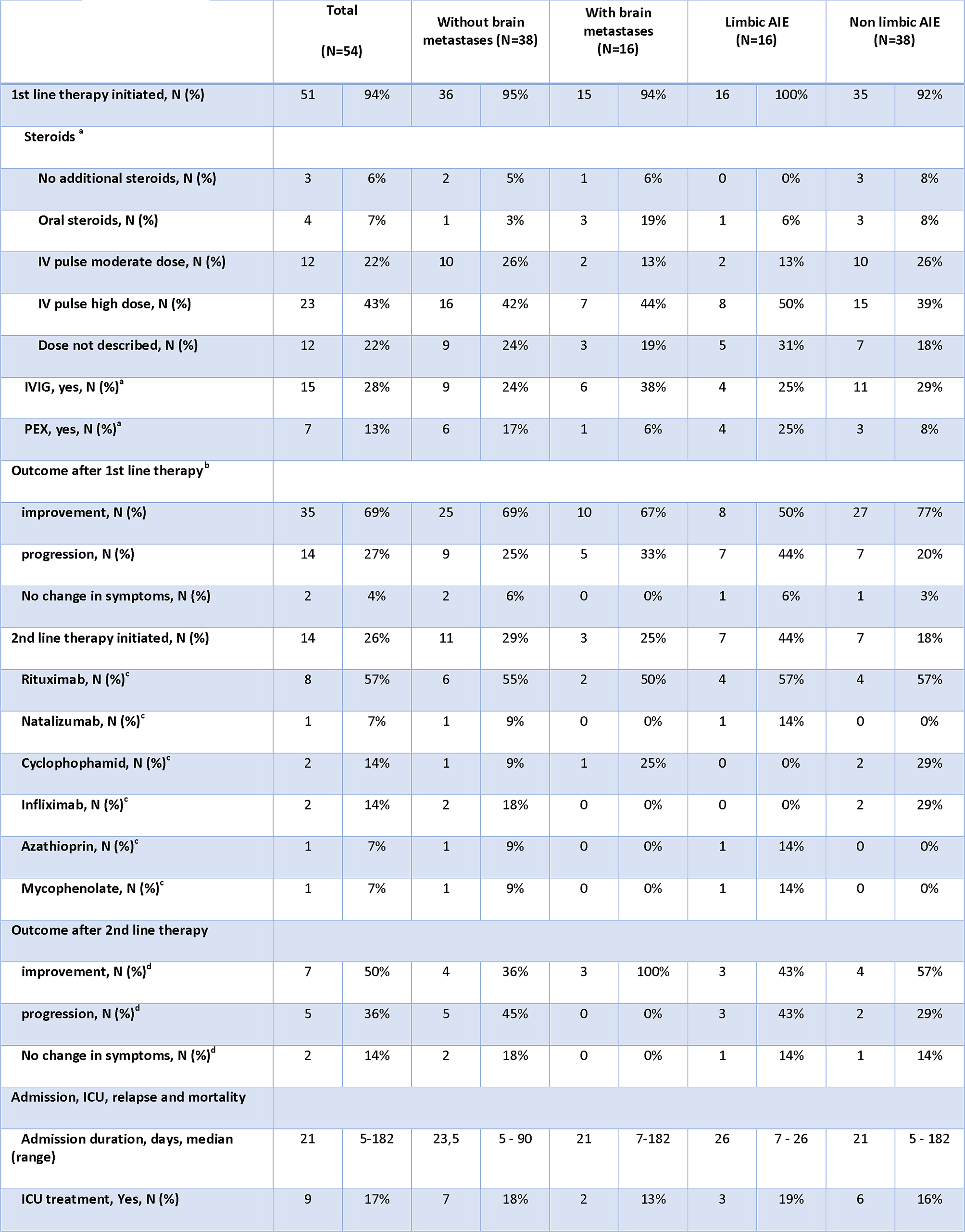
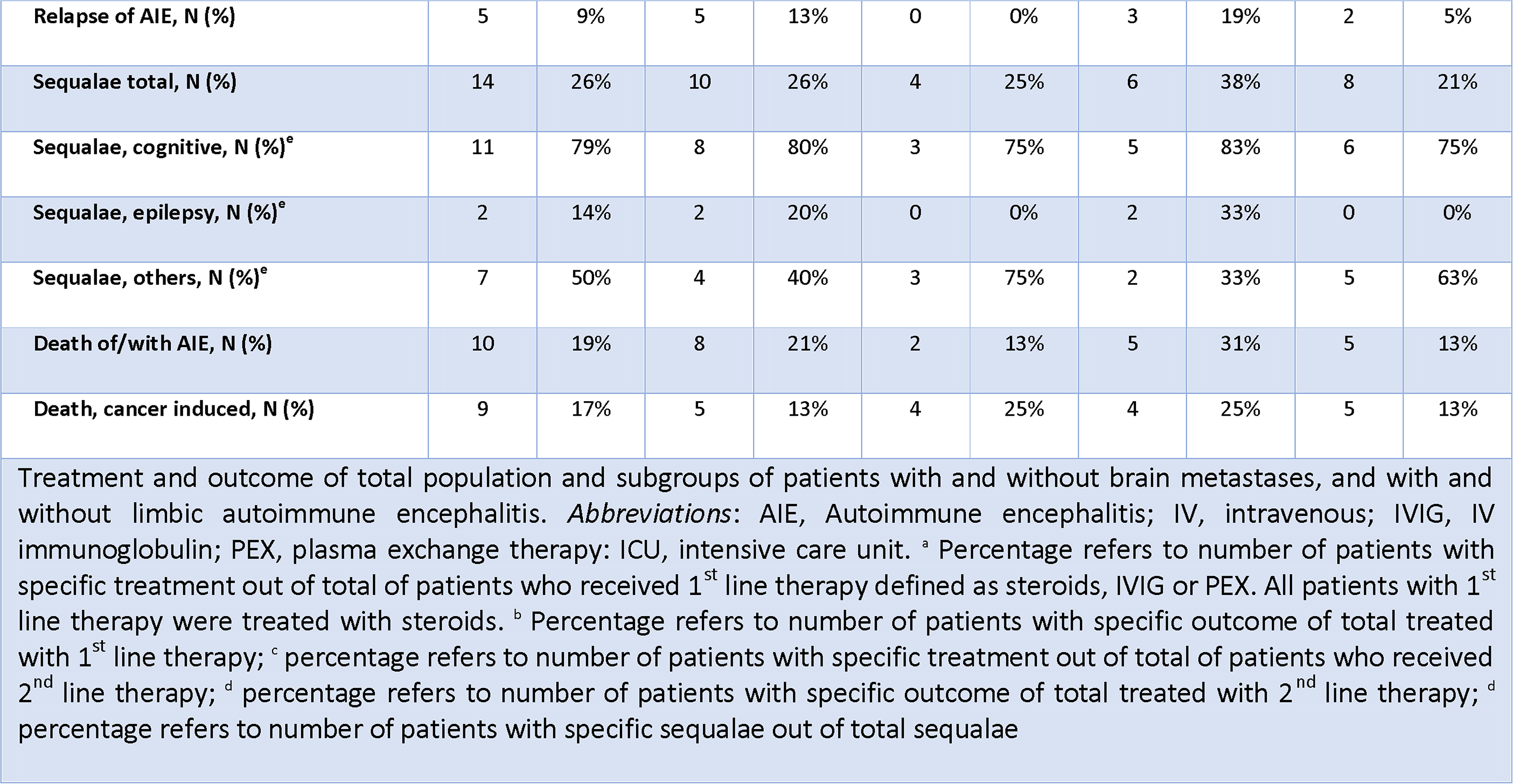

#### First line therapy and outcome

First line therapy was defined as either steroid, IVIG and/or PEX treatment. Steroid treatment was initiated in 51 patients (94%), while one patient did not receive any treatment^12,13^ and two patients were already on steroid treatment which was not increased. Four patients^12,31^ were only treated with oral steroids,^22,35,41,47^ 35 patients received iv pulse therapy of which 23 were given high dose treatment (> 500mg daily), and in 12 patients (22%) steroid dosage was not disclosed. IVIG was initiated in 15 patients (28%). One patient received repeated IVIG treatment.^29^ PEX was initiated in 7 patients^15,18,34,42,46^, including the patient in Figure 1 (13%). All patients treated with either IVIG or PEX, received concomitant steroid treatment. Only two patients received steroids, IVIG and PEX treatment.^18,42^ Improvement after first line therapy was seen in 35 patients (69%), progression of symptoms in 14 patients (27%), and 2 patients (4%) had no change in symptoms after first line therapy. Nine out of 12 patients with progression were started on second line therapy.

#### Second line therapy and outcome

Second line therapy was defined as immunosuppressive agents other than steroids, IVIG or PEX, and was initiated in 14 patients (26%). Eight patients were treated with rituximab,^26,28,31,36,42,46^ one patient with natalizumab,^27^ two patient with cyclophosphamide,^31,45^ two patients with infliximab,^25,30^ one patient with mycophenolate,^19^ and one patient with azathioprine (figure 1). One patient received both rituximab and cyclophosphamide.^31^

Improvement after second line therapy was seen in seven patients ^25–28,31,42^, including the patient in Figure 1, progression in five patients^26,30,36,42,19,45^, and no change in symptoms in two patients.

#### Admission duration, intensive care, relapse and mortality

Data on the length of admission was available for 25 patients (46%) and median of days admitted was 21 (range 5-182). Treatment in an intensive care unit setting was described in nine patients^31–34,26,40,44,48^, including the patient in Figure 1 (17%), and five patients^18,19,25,26,36^ (9%) experienced a relapse of AIE.

Sequalae after admission was described in 14 patients: Prolonged cognitive deficits were most frequent (79%). Death occurred in 19 patients (35%) of which 10 patients^13,26,31,34,36,42,46^ died with/of their presumed AIE, and in nine patients^12,17,18,31,33,39,47^, including the patient in Figure 1, death was attributed to progression of their underlying cancer.

### F. Diagnosis of autoimmune encephalitis

Following application of the AIE criteria proposed by Graus et al.,^7^ 23 patients^11,17,32,33,35,37,38,40,44,46,20–23,28–31^ fulfilled the criteria for possible AIE; 16 patients^14,18,43,46,47,19,22,26,27,34,36,39,42^, including the one in Figure 1, criteria for limbic AIE; two patients^31,41^ criteria for definite anti-NMDA AIE; two patients^10,45^ criteria for probable auto-antibody negative AIE, one patient^16^ criteria for Hashimoto’s encephalopathy (aka. steroid responsive encephalopathy associated with autoimmune thyroiditis, SREAT); and one patient^48^ met criteria for definite acute disseminated encephalomyelitis (ADEM).

Adhering strictly to the proposed criteria, we found nine patients^12,13,15,24–26,32,42,46^ who did not meet the criteria for the following reasons: Four patients^12,13,15,46^ presented with cognitive symptoms of either altered mental status, psychiatric symptoms or working memory deficits but no seizures, focal CNS findings or paraclinical signs of CNS inflammation (although one^32^ showed non-infectious encephalitis with brainstem involvement on post-mortem autopsy). Two patients^24,26^ had focal CNS findings and CSF pleocytosis but no altered mental status, psychiatric symptoms or working memory deficits. One patient^42^ had focal hemiballismus and MRI changes of the basal ganglia but no altered mental status, psychiatric symptoms or working memory deficits. One patient^25^ presented with headache and flu-like symptoms with meningeal enhancement on MRI suggestive of aseptic meningitis. One patient^46^ was described with ophthalmoplegia and head-drop suggestive of affection of the peripheral nerve system without cognitive or any other symptoms suggesting limbic involvement, although MRI showed bitemporal FLAIR lesions.

**Table 6** presents paraclinical findings and outcome in the population divided in subgroups according to the proposed criteria by Graus et al.^7^ A logistic regression was performed to assess if age, sex, presence of brain metastases, presence of intracellular antibodies in CSF and grouping according to Graus et al.^7^ criteria had an effect on improvement after 1^st^ line therapy. We found that the presence of intracellular antibodies was a significant predictor for lack of improvement after 1^st^ line therapy (p < 0.05), but the other variables were non-significant.

**Table 6.**
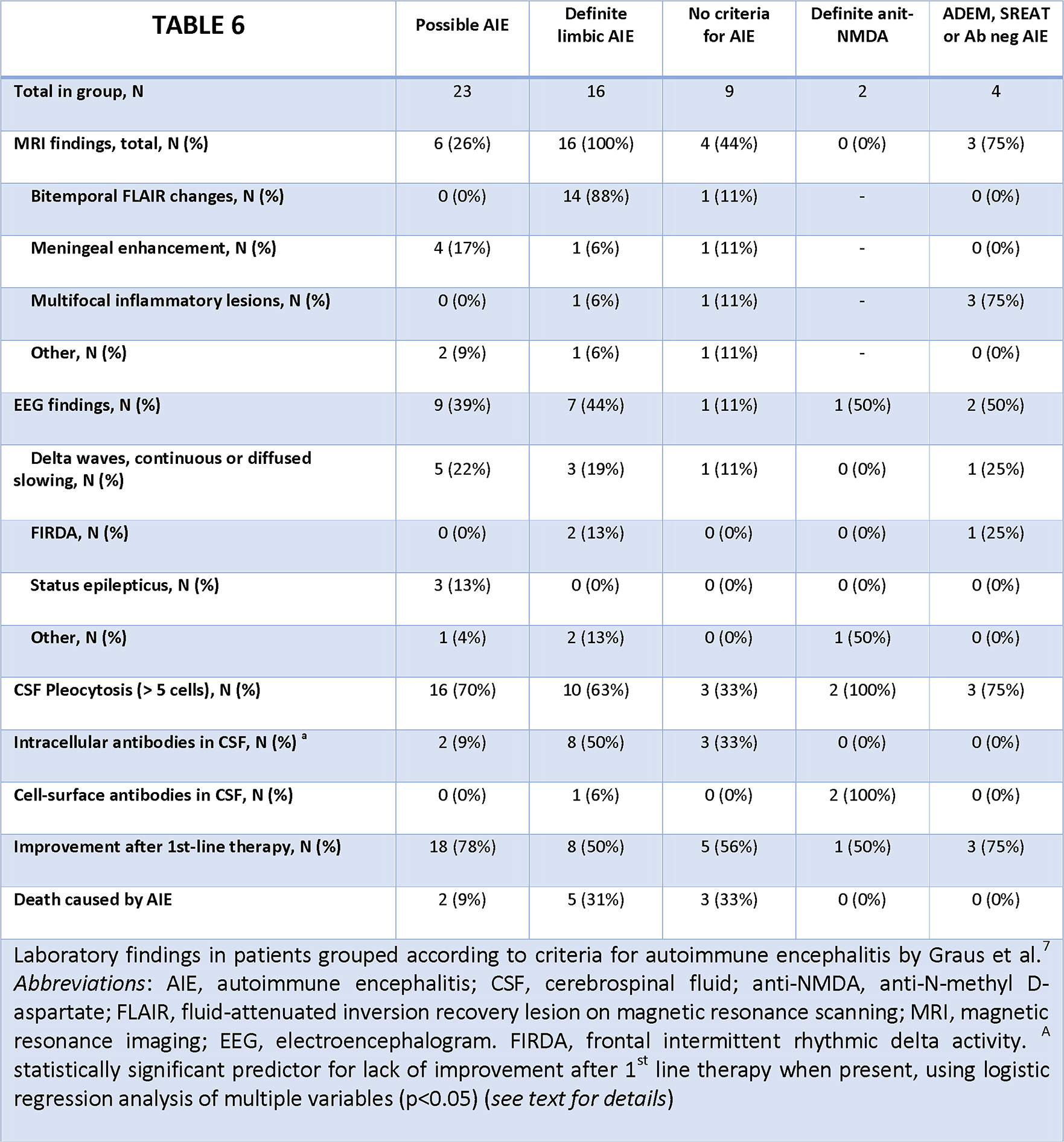

## DISCUSSION

We collected all cases (n=54) of ICPI-associated AIE published since the first description of the entity in 2015 until January 2020. The high number of case reports and case series that have emerged within this comparatively short time period, as well as the wide geographic distribution with reports from the Americas, Europe and Australasia, suggests that ICPI-AIE is a widespread and relevant disorder that neurologists are increasingly likely to encounter in clinical practice.

Several key messages are available for the clinician from this review. First, in patients treated with ICPI for disseminated malignancies, both non-limbic and limbic AIE may arise, but non-limbic AIE presentation is roughly twice as common as limbic AIE. Second, only roughly half of the patients tested have known neuronal antibodies, either in CSF or in blood; but of note, classical intracellular antibodies are more frequent than surface antibodies, perhaps reflecting the paraneoplastic nature of ICPI-associated AIE. Third, AIE criteria as recently proposed by Graus et al.^7^ are applicable to most cases of ICPI-AIE, but we identified 9 of 54 patients who did not meet these criteria for a variety of reasons, so clinical acumen is needed to make a diagnosis of AIE in ICPI treated patients.

Also, the clinical outcome after ICPI-AIE ranged from mild to severe, including the need f or intensive care management and death. Of note, presence of intracellular antibodies was a significant predictor for lack of improvement after 1^st^ line therapy. Overall, however, the underlying cancer is probably the most crucial determinant of survival and morbidity.

Other clinically important messages include the male-female ratio, which was roughly 1:1, so ICPI-associated AIE is unlikely to be sex-specific. Importantly, brain metastases were found in 16 patients (30%), and the presence of cancer spread to the CNS may pose a particular diagnostic challenge as the clinician must decide which signs and symptoms are attributable to AIE and which to the tumor. Also, it must be kept in mind that the average cycle of ICPI treatments before occurrence of symptoms of presumed AIE was 3.5, but very early and very late presentations are possible, and the reasons for this variance are unknown.

Altered mental status appears to be the most frequent presentation in ICPI-AIE, and seizures less so, but seizures and other findings might be underreported given the retrospective nature of the present study. Working memory deficits were more frequent in patients with limbic AIE than in patients with the extra-limbic AIE, which may be explained with the prominent hippocampal involvement in limbic AIE. Laboratory abnormalities comprised a wide range from normal to abnormal. Most frequent were EEG changes, including localized and generalized and epileptiform and non-epileptiform activity, but none of this is specific to ICPI-AIE. CSF pleocytosis appeared equally common in limbic and extra-limbic AIE, but the cell count was less in limbic AIE perhaps reflecting more restricted inflammation. ICPI-associated AIE was most frequent with nivolumab but it is unknown whether nivolumab is particularly prone to inducing AIE or if this simply reflects the more widespread use of it; and it is important to be aware of that AIE may arise with various ICPI, including various combinations of ICPI. Finally, given that ICPI is associated with several AIE presentations, including limbic, non-limbic, SREAT and ADEM, this suggests that the immunological adverse events of ICPI can affect both gray and white matter in different brain regions. This tendency to involve various anatomical sites mirrors what is known of ICPI adverse events affecting the peripheral nervous system; here too, all structures can potentially be involved, including peripheral neurons, neuromuscular junctions and muscle cells.^6^

Reviewing the current data raises important questions, not the least about the pathophysiology of ICPI-associated AIE. For instance, why do some patients experience symptoms after just a single dose of ICPI, while others go through several cycles before AIE onset? Why do some patients develop limbic AIE, while others (the majority) have extra-limbic AIE? What are the molecular targets in antibody-negative patients with atypical presentations or laboratory findings? Exactly how much of the morbidity and mortality are due to AIE and how much to the cancer itself? What is the most appropriate treatment of ICPI-AIE, and should ICPI be discontinued? Future studies are needed to define mechanism and best treatment approaches, but for the time being it seems fair to conclude that neurologists need a high clinical suspicion to identify patients with ICPI-AIE as treatment delay is likely to lead to increased morbidity and mortality.

Systematic reviews like this are prone to certain limitations that should be acknowledged. We may have underestimated the true frequency of AIE characteristics, given that absence of reported signs and symptoms in individual case reports is not necessarily evidence for their factual absence. Consequently, we cannot conclude with confidence if diagnostic inconsistencies are due to inadequate workup or misinterpretation of results. Further, we were unable to quantify treatment effects and mortality compared with non-ICPI AIE because the data lacked sufficient detail and direct comparisons between ICPI-AIE and non-ICPI-AIE are unavailable. Moreover, the effects of metastatic cancer itself may be difficult to distinguish from severe autoimmune disease, requiring further study.

To summarize, the high number of reports on ICPI-AIE that have emerged within the past 5 years suggests that neurologists are increasingly likely to see this disorder in their daily practice. While both limbic and non-limbic AIE presentations are common, the latter is even more frequent. Intraneuronal antibodies are more common than neuronal surface antibodies, and data indicate that presence of intraneuronal antibodies may negatively affect AIE treatment response. Finally, many important questions remain, in particular related to pathophysiological mechanisms, clinical symptomatology and treatment. Thus, autopsy studies are needed to define mechanisms; prospective registry studies are required to define prevalence; and randomized clinical trials are important to establish the best treatment options in ICPI-associated AIE.

## Data Availability

Not applicable - all relevant data are presented in the Tables and the main manuscript body

## Financial Disclosures

The authors have no disclosures to declare.

## Study funding

None

## Patient consent

Patient consent for Figure 1 was obtained.

This review was pre-registered at PROSPERO: https://www.crd.york.ac.uk/prospero/display_record.php?RecordID=139838.

---

**Figure S1.**
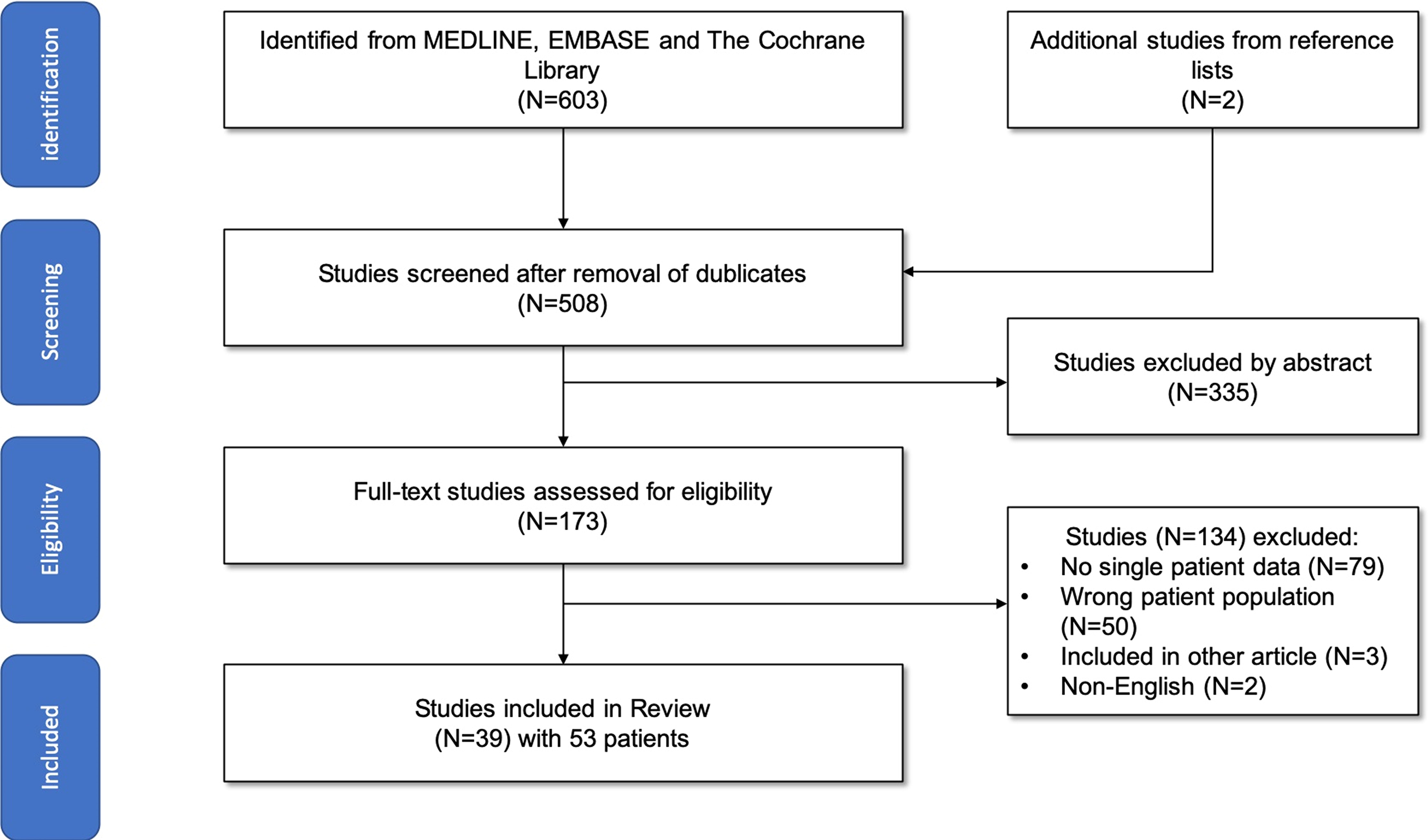
Flowchart of the literature search

## Notes

### Competing Interest Statement

The authors have declared no competing interest.

### Author Declarations

This is a systematic review, so no IRB required

